# Review and methodological analysis of trials currently testing treatment and prevention options for COVID-19 globally

**DOI:** 10.1101/2020.04.27.20080226

**Authors:** Paraskevi C. Fragkou, Drifa Belhadi, Nathan Peiffer-Smadja, Charalampos D. Moschopoulos, François-Xavier Lescure, Hannah Janocha, Emmanouil Karofylakis, Yazdan Yazdanpanah, France Mentré, Chrysanthi Skevaki, Cédric Laouénan, Sotirios Tsiodras, on behalf of the ESCMID Study Group for Respiratory Viruses.

## Abstract

**Background:** As COVID-19 cases continue to rise globally within an unprecedented short period of time, solid evidence from large randomised controlled trials is still lacking. Currently, numerous trials testing potential treatment and preventative options are undertaken globally.

**Objectives:** We summarised all currently registered clinical trials examining treatment and prevention options for COVID-19. Additionally, we evaluated the quality of the retrieved interventional studies.

**Data sources:** Clinicaltrials.gov, the Chinese Clinical Trial Registry and the European Union Clinical Trials Register were systematically searched.

**Study eligibility criteria:** Registered clinical trials examining treatment and/or prevention options for COVID-19 were included. No language, country or study design restrictions were applied. We excluded withdrawn or cancelled studies and trials not reporting therapeutic or preventative strategies for COVID-19.

**Participants and interventions:** No restrictions in terms of participants’ age and medical background or type of intervention were enforced.

**Methods:** The registries were searched using the term “coronavirus” or “COVID-19” from their inception until 26^th^ March 2020. Additional manual search of the registries was also performed. Eligible studies were summarised and tabulated. Interventional trials were methodologically analysed, excluding expanded access studies and trials testing Traditional Chinese Medicine.

**Results:** In total, 309 trials evaluating therapeutic management options, 23 studies assessing preventive strategies and 3 studies examining both were retrieved. Interventional treatment studies were mostly randomised (n=150, 76%) and open-label (n=73, 37%) with a median number of planned inclusions of 90 (IQR 40-200). Major categories of interventions that are currently being investigated are discussed.

**Conclusion:** Numerous clinical trials have been registered since the onset of the COVID-19 pandemic. Summarised data on these trials will assist physicians and researchers to promote patient care and guide future research efforts for COVID-19 pandemic containment. However, up to the end of March, 2020, significant information on reported trials was often lacking.

## INTRODUCTION

COVID-19 pandemic induced by the Severe Acute Respiratory Syndrome coronavirus 2 (SARS-CoV-2) has already affected most countries globally [1]. As of April 12, 2020, more than 1,690,000 COVID-19 cases have been officially reported leading to more than 150,900 deaths worldwide, with numbers constantly rising on a daily basis as the virus continues to spread exponentially [2,3].

The original source of SARS-CoV-2 is still debatable, though phylogenetic data analysis converge into the hypothesis that SARS-CoV-2 has possibly originated from bat SARS-like coronaviruses (CoVs) [4–6]. Human CoVs are positive-sense single-stranded RNA (+ssRNA) viruses belonging to the *Orthocoronavirinae* subfamily [7]. The analysis of viral genome as well as structural and non-structural proteins plays central role in identifying candidate treatment targets [7]. Notably, the SARS-CoV-2 surface spike glycoprotein (S-protein) is a key structural component that controls the viral interaction with hosts’ angiotensin I converting enzyme 2 (ACE2) receptor in order to invade human epithelial cells [8]. Moreover, molecules directed against non-structural proteins such as the viral protease or pathways imperative for viral genome replication, alone or along with boosters or regulators of the host’s immune system, constitute promising therapeutic strategies[8,9].

Given the steep upsurge of COVID-19 cases worldwide within an unprecedented short period of time, we are still waiting for solid evidence from large randomised controlled trials regarding targeted antiviral treatments. In this systematic review, we aim firstly to summarise the data on all currently tested treatment and prevention options for the COVID-19 disease, and secondly to methodologically analyse and evaluate the quality of the registered interventional studies.

## METHODS

Registered clinical trials were systematically searched at the ClinicalTrials.gov database[10], the Chinese Clinical Trial Registry[11] and the European Union Clinical Trials Register [12] from their inception up to 26^th^ March 2020 using the search terms “coronavirus” or “COVID-19”. Additional manual search of the registries was performed for possibly unidentified studies. No language, country or study design restrictions were applied. Participants of any age and medical background with or at risk for COVID-19 were included, as were any currently tested intervention related to the treatment or prevention of COVID-19.

We excluded withdrawn or cancelled studies and trials not reporting therapeutic or preventative measures for COVID-19. Eligible studies were summarised and tabulated.

Methodological analysis of the interventional studies was performed. Traditional Chinese Medicine (TCM) and homeopathy were excluded from the in-depth qualitative assessment, as we have no expertise to analyse clinical trials testing these agents that rely on controversial scientific rationale [13,14]. We evaluated the study design, number of planned inclusions and primary outcomes of interventional studies, excluding retrospective and “expanded access” studies. Studies were also analysed according to their primary endpoint, i.e. clinical, virological (viral excretion in clinical samples), radiological (imaging results such as CT-scan or X-rays), or biological / immunological (complete blood count, CR, CD8+/CD4+ T cells count, etc.). This review was conducted according to the PRISMA guidelines [15].

## RESULTS

### 1. General data retrieved

In total, 335 studies were retrieved, including 309 trials evaluating therapeutic molecules, devices and other management options, 23 studies assessing medications, vaccines under development and other preventive strategies, and 3 studies examining both (Figure 1, Table S1, Table S2). An overview of currently tested therapeutic interventions is presented in Figure 2 and Figure 3.

**Figure 1.**
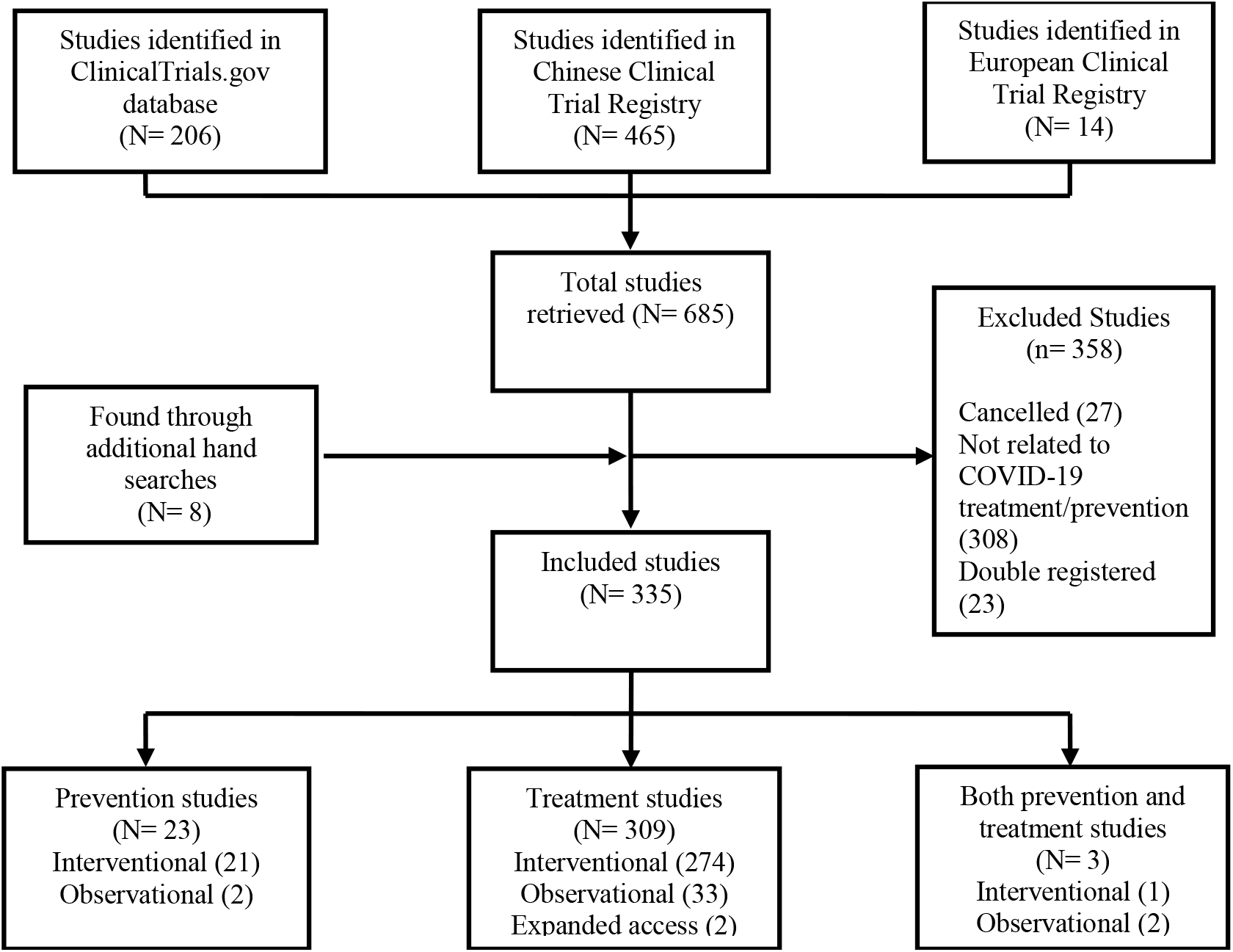
Systematic review Flow Chart.

**Figure 2.**
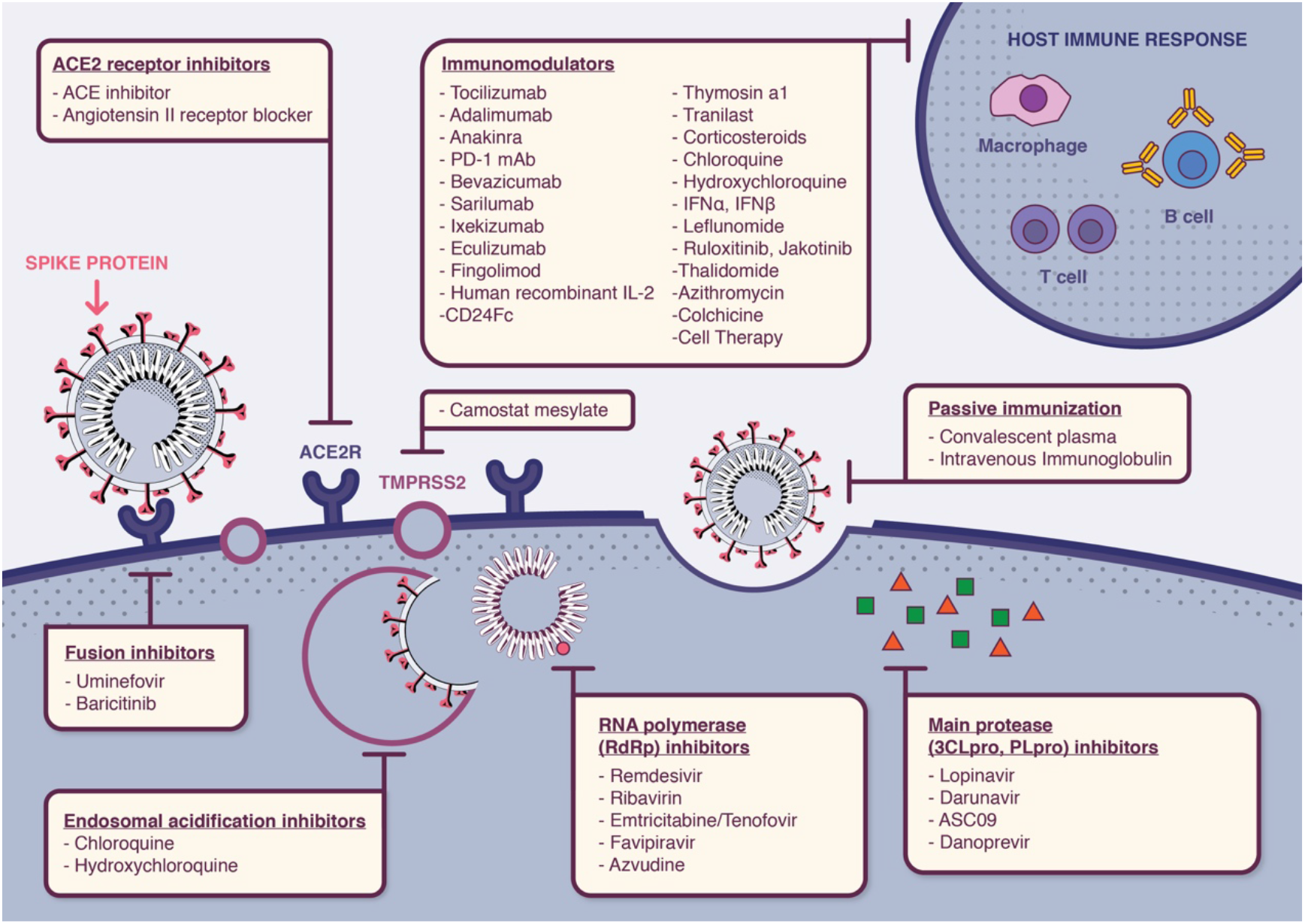
Currently tested therapeutic molecules targeting different steps of SARS-CoV-2 life cycle.

**Figure 3.**
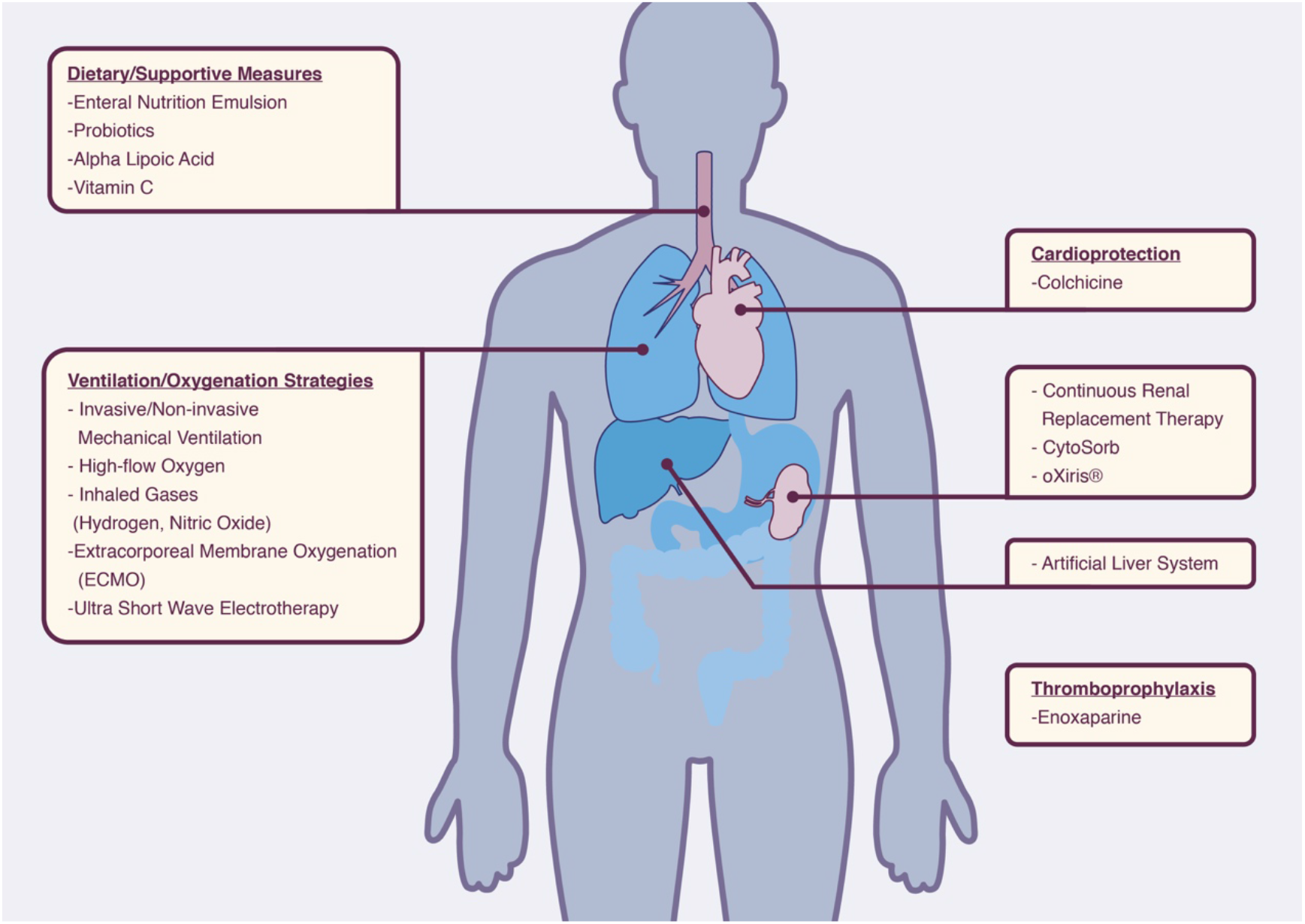
General and supportive therapeutic interventions tested for novel coronavirus disease (COVID-19).

### 2. Main treatment interventions

#### Protease Inhibitors

Lopinavir is a protease inhibitor (PI) activeagainst human immunodeficiency virus 1 (HIV-1) infection. The main coronavirus proteinase (3C-like proteinase or 3CL^pro^) plays a key role in processing viral polyproteins [16,17]. PIs effectively inhibit the 3CL^pro^ enzyme, thus posing a possibly potent therapeutic agent against SARS-CoV-2 infection.

PIs have shown effectiveness against SARS-CoV, MERS-CoV and SARS-CoV-2 viruses in *in vitro* susceptibility models [18–21]. During SARS-CoV epidemic, lopinavir boosted by ritonavir (a cytochrome P450-3A4 inhibitor) with or without ribavirin, significantly reduced adverse outcomes including mortality[22]. The MIRACLE trial that examines the efficacy of ritonavir boosted lopinavir combined with recombinant interferon-beta 1b (IFN-β1b) in the treatment of MERS, is currently undertaken in Saudi Arabia and results are pending [23].

For COVID-19, lopinavir/ritonavir combined with or without other agents has been reported to successfully reduce adverse outcomes in sporadic cases from China [24–27]. These promising reports have set the ground for numerous trials addressing the safety and efficacy of PIs in SARS-CoV-2 infection (Table 1). Other PIs that are currently being assessed are ritonavir boosted ASC09 (a novel PI), cobicistat boosted darunavir as well as the NS3/4A protease inhibitor danoprevir combined with ritonavir (Table 1, Table S1).

**Table 1.**
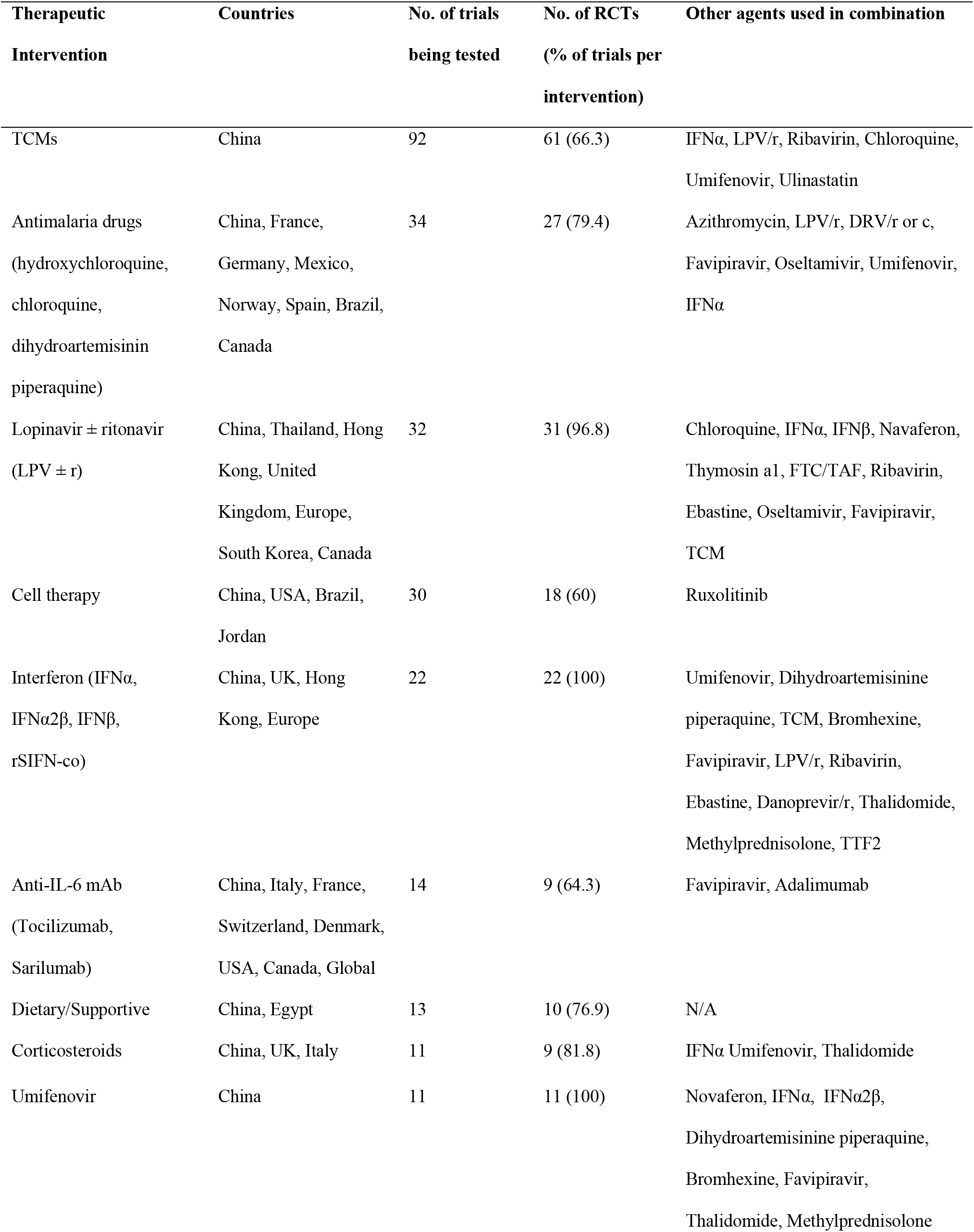
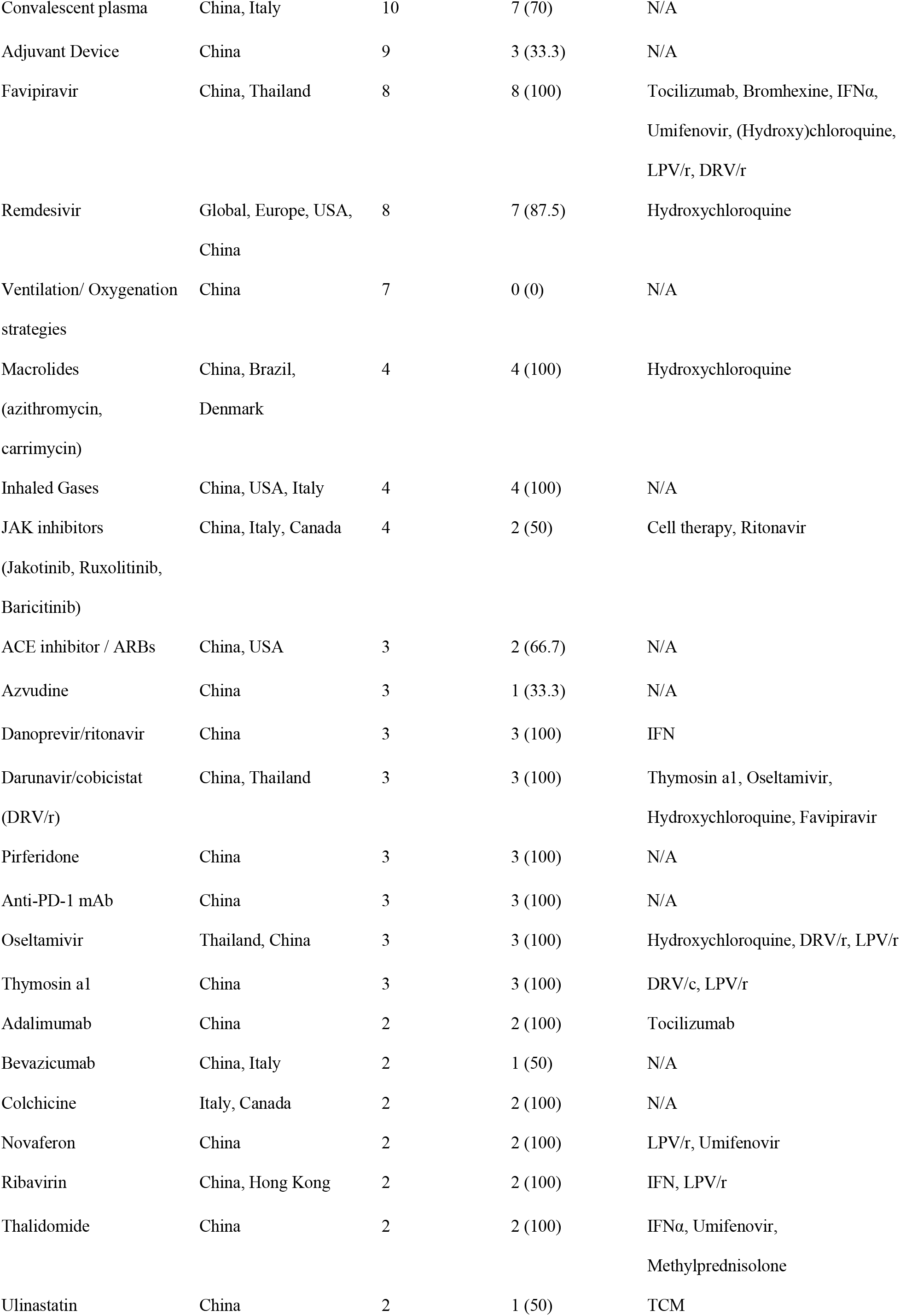
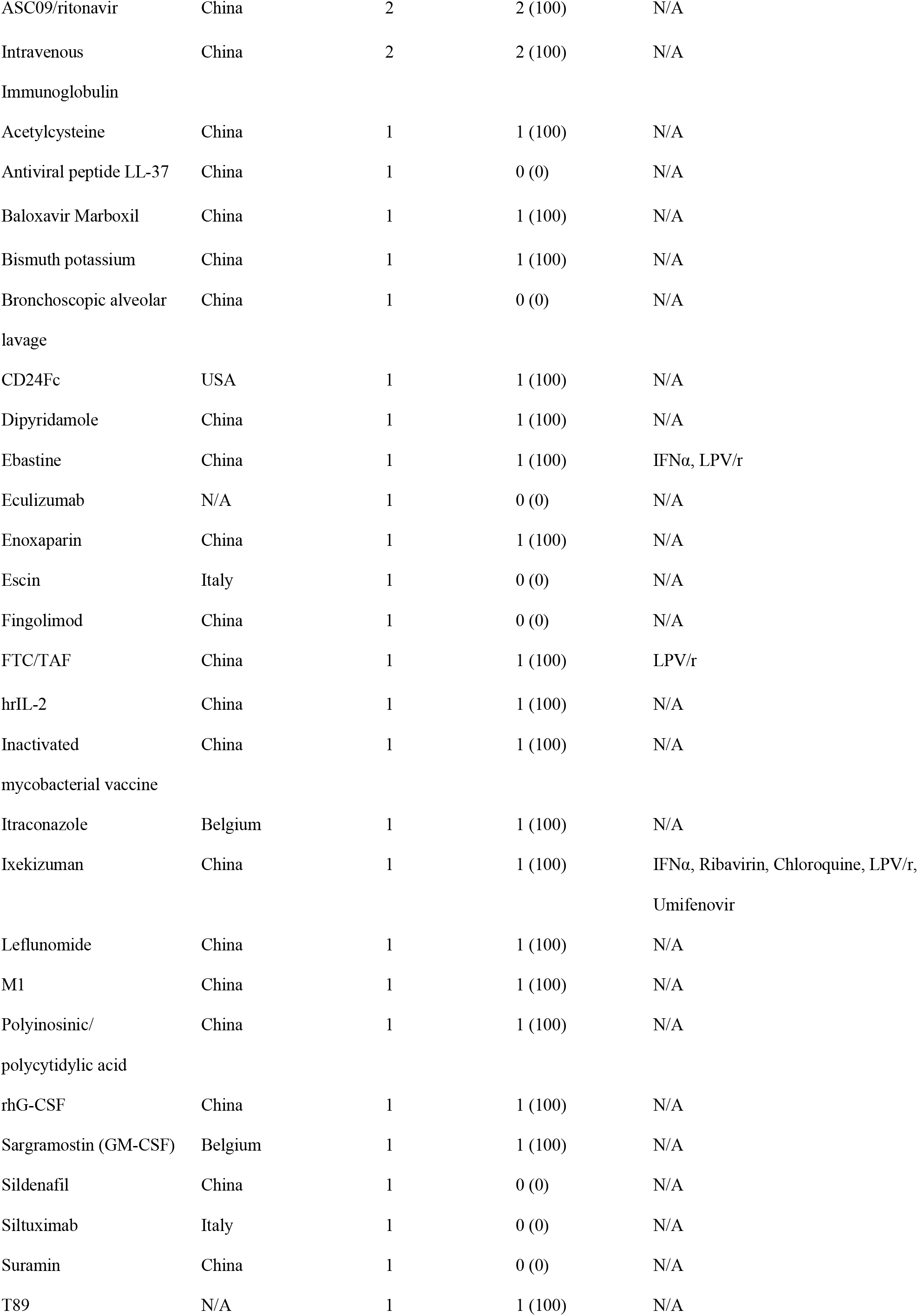
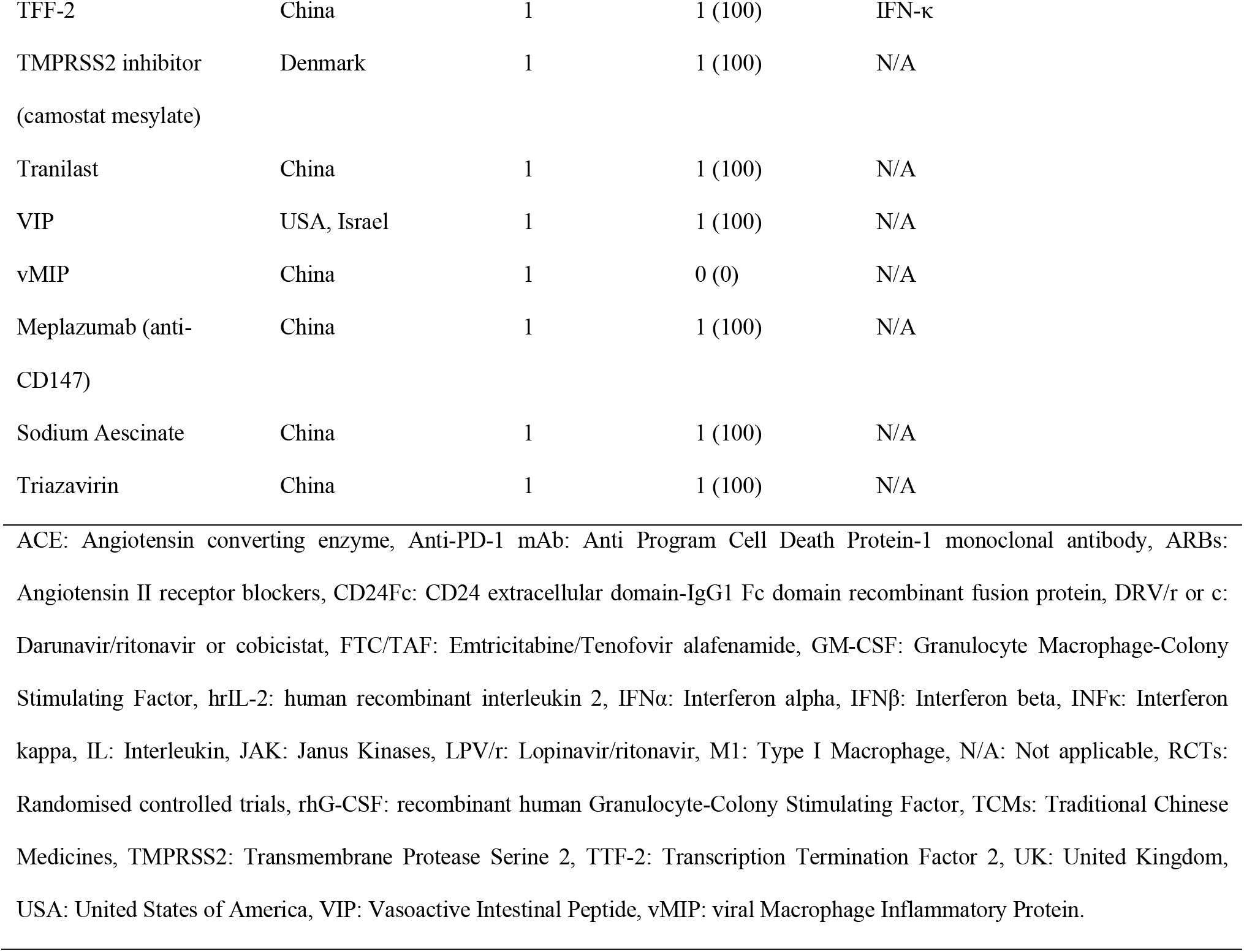
Treatment interventions currently being evaluated for the novel coronavirus disease (COVID-19) globally.

#### RNA polymerase inhibitors

SARS-CoV-2 and SARS-CoV RNA-dependent RNA polymerase (RdRp) share 96% sequence identity; this has justified the assumption that inhibitors effective against SARS-CoV could have similar inhibitory effects against SARS-CoV-2[28]. Nucleoside analogues compete with natural nucleosides for the RdRp active site, thus inhibiting the viral genome replication [29]. Current research efforts focus on repurposing older moleculesinCOVID-19 treatment, as their safety profile has already been documented [30].

Remdesivir (GS-5734^TM^) is an adenosine analogue with broad-spectrum antiviral properties [31]. Preclinical data demonstrated the efficacy of remdesivir against SARS-CoV and MERS-CoV, as it has the potential to outcompete the proofreading ability of coronavirus exonuclease, and carries a high genetic resistance barrier [21,28,30,32]. Wang et al. confirmed its *in vitro* efficacy against SARS-CoV-2 [33].Currently, it is investigated in 7 randomised, controlled trials (Table 1, Table S1).

Favipiravir is a nucleoside analogue inhibiting the RNA polymerase, initially approved for the treatment of novel influenza viruses [34]. It is also effective against a broad range of viruses, including positive-sense single-stranded RNA viruses [34]. Since there have been some promising *in vitro* results for its efficacy against SARS-CoV-2, favipiravir is now being investigated in 8 clinical trials.

Ribavirin is a guanosine analogue that inhibits inosine monophosphate dehydrogenase required for the synthesis of guanosine triphosphate, leading to lethal mutagenesis of RNA genome [35]. Ribavirin was used in SARS epidemic in combination with either lopinavir/ritonavir or interferon alpha (IFN-α), and these combinations are currently recommended by the China National Practice Guidelines for the treatment of severe COVID-19 [22,36].

Azvudine, an azidocytidine analogue that inhibits viral reverse transcriptase,has been effective against HIV, hepatitis B and C viruses [37]. Its efficacy against SARS-CoV-2 is being tested in 3 ongoing clinical trials (Table 1, Table S1). Another nucleoside analogue undergoing investigation for COVID-19 pneumonia is emtricitabine/tenofovir alafenamide.

#### Anti-malaria drugs

Chloroquine and hydroxychloroquine are currently licensed for the treatment of malaria and autoimmune diseases [38]. However, they have also been studied against several viruses with promising *in vitro* results, never confirmed in humans [39–41]. As weak bases, they are concentrated in acidic intra-cellular organelles, leading to alkalization and distraction of the low pH-dependent steps of viral replication, including viral-cell fusion and uncoating [38,40]. Moreover, they impair the terminal glycosylation of ACE2 receptor in Golgi apparatus, thus inhibiting the viral penetration into the host cells [42].

As they are accumulated in lymphocytes and macrophages, these drugs reduce secretion of proinflammatory cytokines, and particularly of tumour necrosis factor alpha (TNF-α) [41]. Experimental data demonstrated that chloroquine is highly effective *in vitro* against SARS-CoV-2 in an estimated effective concentration that is easily achievable with standard dosing regimens [33]. Preliminary results from small studies suggested the effectiveness of chloroquine, alone or combined with azithromycin, in viral clearance, inhibition of pneumonia exacerbation, improvement of lung imaging and shortening of the disease course [43,44]. However, clinical trials with a control group are needed to provide reliable answers for clinicians.

#### Immunomodulators and anti-inflammatory drugs

Virus-induced exuberant immune response leading to cytokine storm syndrome (CSS) and secondary haemophagocytic lymphohistiocytosis (HLH) is probably the underlying pathogenetic mechanism that leads to critical and often fatal COVID-19 infection [45,46]. Hyperinflammation is associated with acute respiratory distress syndrome (ARDS) and fulminant multi-organ failure thatarefatal if left untreated. In this context, immunosuppressors (along with antivirals) may play a key role in counteracting severe SARS-CoV-2 infection [45].

Preliminary data from COVID-19 patients in China reported significantly higher interleukin (IL)-6 levels in patients with critical COVID-19 disease than those with severe or mild disease [47]. Tocilizumab and sarilumab, both humanised monoclonal antibodies (mAbs) against the IL-6 receptor, are currently evaluated in 14 clinical trials (Table 1, Table S1); tocilizumab, in particular, improved symptoms and laboratory parameters in a small retrospective study in China [48]. Various other molecules oriented against different cytokines, as well as intra-and extra-cellular inflammatory pathways, are currently being tested in COVID-19 including: adalimumab, anti-programmed cell death protein 1 mAbs, bevacizumab, ixekizumab, eculizumab, human recombinant IL-2, inhibitors of NLRP3 inflammasome activation (tranilast), Janus Kinase inhibitors, fingolimod and a recombinant fusion protein targeting an immune pathway checkpoint (CD24Fc).Clinical trials on other immunotherapies (i.e.gimsilumab, siltuximab, lenzilumab and leronlimab) have been proposed, but havenot been registered yet.

Immunomodulators licensed for haematological and rheumatological conditions (such as leflunomide and thalidomide), as well as colchicine that counteracts the assembly of the NLRP3 inflammasome, are also being studied for their therapeutic use against SARS-CoV-2 (Table 1, Table S1) [49].

The immunomodulatory effects of macrolide antibiotics, as well as their pharmacodynamic property to achieve at least 10-fold higher concentrations in epithelial lung fluid than in serum, have led researchers to repurpose them against SARS-CoV-2 (Table 1, Table S1) [50,51]. Preliminary results of a study have shownsignificant viral load reduction in patients treated withhydroxychloroquine plus azithromycin [43].

Corticosteroids are major anti-inflammatory drugs, with a conflicting safety profile in severe viral infections [52]. However, their useis currently recommended by the European Society of Intensive Care Medicine in COVID-19 cases with shock and/or evidence of CSS/HLH and/or mechanically ventilated patients with ARDS [53]. Corticosteroids are being tested in 11 clinical trials (Table 1, Table S1).

Finally, immunostimulatory molecules that enhance the hosts’ immune response against the invading pathogen, like IFN-α, interferon beta (IFN-β), the recombinant protein produced by DNA-shuffling of IFN-α (novaferon), the inactivated *Mycobacterium* vaccine and thymosin a1, are evaluated as viable therapeutic options in various combinations against SARS-CoV-2 [54].

#### Membrane fusion inhibitors and inhibitors of ACE2 receptor connection

SARS-CoV-2S-protein bindsACE2 receptor on the epithelial cells’ membrane [55]. The host’s type II transmembrane serine protease (TMPRSS2) facilitates membrane fusion and augments viral internalization by cleaving and activating the S-protein [56,57]. These proteins, being an integral part of the viral life cycle, can be used as possible therapeutic targets.

ACE inhibitors (ACEi) selectively inhibit ACE1 but not ACE2, as they are structurally different enzymes [58]. Experimental animal models show that angiotensin II type 1 receptor blockers (ARBs) and ACEi may up-regulate ACE2 expression; although this raised concerns that ACEi and ARBs may be associated with a more severe form of COVID-19, there is no evidence supporting that ACE2 up-regulation augments the virulence or the penetration of the virus into the hosts’ cells as of yet [58,59]. Contrarily, renin-angiotensin system (RAS) inhibition may ameliorate the over-accumulation of angiotensin II induced by the down-regulation of ACE2 noted in other CoVs infection, thus possibly protecting against the development of fulminant myocarditis and ARDS [58]. In spite of controversial data, 2 randomised controlled trialstesting losartan are currently undertaken in the USA.

Camostat mesilate is a potent serine protease inhibitor, inhibiting the TMPRSS2 and is approved for chronic pancreatitis [60]. *In vitro* experiments have shown that camostat is effective against SARS-CoV-2[61]. Nafamostat, another inhibitor of TMPRSS2 has been suggested in COVID-19 therapy, but no registered trials are available yet.

Umifenovir is a small indole derivative with a broad-spectrum antiviral activity developed about 30 years ago [62,63]. Although it is currently used in Russia and China to combat influenza, it has shown *in vitro* activity against numerous DNA and RNA viruses including SARS-CoV [62,64,65]. It primarily targets the interaction between viral capsid and the membrane of the host cell by inhibiting the attachment, the fusion and the internalization of the virus [62]. Moreover, umifenovir has direct virucidal effects against enveloped viruses through interaction with the viral lipid envelope or with key residues within structural proteins of virions [62,65,66]. Limited data support that umifenovir may exhibit immunomodulatory effects by stimulating hosts’ humoral immune response, interferon production, and phagocytes’ activation [67]. A small retrospective study in China demonstrated some promising results regarding its effectiveness against SARS-CoV-2 when used in combination with lopinavir/ritonavir [68]. Currently 11 relevant clinical trials are in progress globally (Table 1, Table S1).

#### Passive immunization

The use of convalescent plasma (CP) to passively immunise patients against viral pathogens has been previously reported, especially when no other treatment options wereavailable [69]. CP obtained from recovering patients was usedduringthe SARS epidemic; two retrospective studies demonstrated that CP administration in SARS patients reduced duration of hospitalisation and mortality rates, when treatment with ribavirin and corticosteroids failed [70–72].

A meta-analysis of patients with severe respiratory infection induced by various viruses showed a statistically significant reduction of 75% in mortality odds in those who received CP across all viral aetiologies, including the influenza A 2009 pandemic strain (H1N1pdm09) and SARS-CoV[73]. Evidence of survival benefit was noted after early administration, while no serious adverse effects were reported. Although results are pending from 10 relevant clinical trials, the potential short-lasting immunity after CoV infection has raised uncertaintyregardingthe clinical efficacy of CP antibodies in clinical practice[74,75].

#### Cell therapies

Mesenchymal stromal cells (MSCs) exhibit immunomodulating qualities, may skew immune cell differentiation and have shown promise in H5N1-associated acute lung injury in preclinical models[76]. A pilot study involving MSC transplantation in 7 patients with COVID-19 resulted in cure or significant improvement of pulmonary function and symptoms without adverse effects [77]. MSCs may partially accumulate in lungs and improve the pulmonary microenvironment. A significant induction of regulatory dendritic cells(DCs), along with a shift from Th1 towards Th2 immune responses, was also observed [77]. Other clinical trials utilizing MSCs for curing COVID-19 are ongoing (Table 1, Table S1).

MSC-derived exosomes are extracellular bodies mediating intercellular communication, containing mRNAs, miRNAs, lipids, growth factors and cytokines, that possibly exert the paracrine immunoregulatory effects of MSC [78]. Furthermore, MSC-exosomes’ administration is associated with lower risks (e.g.tumour formation, immunogenicity) compared to the intravenous injection of MSCs [79].

Natural killer cells have shown promising results as adoptive immunotherapy in various cancers, and may also be potent effector cells in the SARS-CoV-2 infection. A phase 1 clinical trial has been launched to determine their efficacy (Table S1). Lastly, ozone therapy may inactivate viruses, among other pathogens, and activate the immune system via up-regulation of Th1 cytokines including IFN and TNF; as such,3 trials have been launched to evaluate ozone autohaemotherapy efficacy in COVID-19 [80].

### 3. Main preventative measures

The World Health Organization (WHO) and the European Centre for Disease Prevention and Control (ECDC) emphasise the role of screening, precaution measures, exposure prevention and environmental disinfection as the mainstay of COVID-19 prevention[81,82]. As no effective preventative options are available yet, several clinical trials are under way to explore the efficacy of various prevention strategies (Table 2, Table S2).

**Table 2.** Prevention interventions currently being evaluated for the novel coronavirus disease (COVID-19) globally.

| Prevention intervention | Countries | No. of trials being tested | No. of RCTs (% of trials per intervention) | Other agents used in combination |
| --- | --- | --- | --- | --- |
| TCMs | China | 10 | 3 (30) | N/A |
| Antimalaria Drugs | China, USA, Mexico, UK, Spain | 6 | 6 (100) | Umifenovir, DRV/c |
| Vaccine | USA, China | 4 | 0 (0) | N/A |
| Health education | China, Hungary | 3 | 2 (66.7) | TCM |
| IFNα1β | China | 2 | 0 (0) | Thymosin α1 |
| Darunavir/cobicistat | Spain | 1 | 1 (100) | Chloroquine |
| Lopinavir/ritonavir | Canada | 1 | 1 (100) | N/A |
| Mask | Canada | 1 | 1 (100) | N/A |
| PUL-042 | USA | 1 | 1 (100) | N/A |
| Thymosin α1 | China | 1 | 0 (0) | IFNα1β |
| Umifenovir | China | 1 | 1 (100) | Hydroxychloroquine |
| DRV/c: Darunavir/cobicistat, IFNα: Interferon alpha, LPV/r: Lopinavir/ritonavir, N/A: Not applicable, RCTs: Randomised controlled trials, TCMs: Traditional Chinese Medicines, UK: United Kingdom, USA: United States of America. | | | | |

Many studies are currently evaluating the efficacy of TCM in COVID-19 prevention in China. Importantly, at least 4 vaccines are under development. Among them, an mRNA-based vaccine encoding the S-protein is being assessed for its safety, reactogenicity and efficacy against SARS-CoV-2 (Table 2, Table S2). Besides the registered trials, other large companies have also announced the initiation of vaccine development [83,84].

Other preventative molecules include hydroxychloroquine and the recombinant human interferon α1b spray. In the USA, exposed individuals are randomised to hydroxychloroquine or placebo, evaluating the agent’spotential as post-exposure prophylaxis (NCT04308668, Table S2). Furthermore, another randomised clinical trial evaluates the efficacy of a 3-month course of chloroquine in at-risk healthcare personnel (NCT04303507, Table S2). Finally, the live attenuated strain of *Mycobacterium bovis* is expected to be tested as a preventative strategy against COVID-19 among healthcare professionals, in Australia and France.

### 4. Methodological analysis of interventional studies

#### Study Populations

In total, 198 interventional treatment and 16prevention trials were included in the methodological analysis respectively (Table 3). Among the eligible treatment studies, children recruitment (i.e.< 14 years old) was reported in 7 clinical trials in total: 1 testing darunavir with cobicistat (NCT04252274); 2 on human stem cells transfusion (ChiCTR2000029606, ChiCTR2000030944); 1 testing hydroxycholoroquine (EudraCT Number: 2020-000890-25); 1 using tocilizumab (NCT04317092); and 1 assessing nutritional supplements (NCT04323345) (Table S1).With respect to relevant prevention studies, children were included in 2 vaccine trials(NCT04276896and NCT04299724) as shown in Table S2.

**Table 3.** Description of the clinical trials registered for the treatment and prevention of COVID-19.

| Study characteristics |  | Treatment studies | Prevention studies |
| --- | --- | --- | --- |
|  |  | N = 198 (%) | N = 16 (%) |
| Study phase |  |  |  |
|  | Phase I | 9 (5) | 3 (19) |
|  | Phase II* | 31 (16) | 2 (13) |
|  | Phase III** | 35 (18) | 6 (38) |
|  | Phase IV | 40 (20) | 1 (6) |
|  | Unspecified | 83 (42) | 4 (25) |
| Study design |  |  |  |
|  | Randomised | 150 (76) | 10 (63) |
|  | Non-randomised | 21 (11) | 3 (19) |
|  | Single-arm | 27 (14) | 3 (19) |
| Blinding |  |  |  |
|  | Double-blind | 31 (16) | 6 (38) |
|  | Single-blind | 16 (8) | 2 (13) |
|  | Open-label | 73 (37) | 5 (31) |
|  | Non-applicable† | 29 (15) | 3 (19) |
|  | Unspecified | 49 (25) | 0 (0) |
| Total number of planned inclusions |  |  |  |
|  | <50 | 62 (31) | 1 (6) |
|  | 50-100 | 41 (21) | 0 (0) |
|  | 100-150 | 32 (16) | 3 (19) |
|  | 150-200 | 9 (5) | 0 (0) |
|  | 200-250 | 15 (8) | 1 (6) |
|  | ≥250 | 39 (20) | 11 (69) |
| Primary endpoint |  |  |  |
|  | Clinical | 128 (65) | 13 (81) |
|  | Virological | 39 (20) | 2 (13) |
|  | Radiological | 19 (10) | 0 (0) |
|  | Immunological/biological | 12 (6) | 1 (6) |

\*Including phase I/II trials; \*\*Including phase II/III trials; †: Single-arm or factorial trials.

#### Study designs

Phase IV and phase III treatment trials were the most commonly reported interventional study types (n=40, 20% and n=35, 18% respectively) as demonstrated in Table 3. Nonetheless, the majority of registered trials do not disclose the study phase (n=83, 42%).

In terms of blinding, 73 open-label (37%), 31 double-blinded (16%), and 16 single-blinded (8%) studies were retrieved. Most trials were randomised (n=150, 76%) with a parallel assignment between arms. The median (IQR) number of planned inclusions is 90 (40-200) with a range of 5 to 6000 participants.

Phase III and phase I prevention studies were the most commonly reported ones (n=6, 38% and n=3, 19% respectively, Table3). As with treatment trials, many prevention trials do not report the study phase (n=4, 25%).

Regarding prevention studies’ blinding, 6double-blinded (38%), 5open-label (31%), and 2 single-blinded (13%) were found. Most studies were randomised (n=10, 63%) with a parallel assignment design. The median (IQR) number of planned inclusions is 513 (177-2958) ranging from 45 to 7576 participants.

#### Treatments and interventions

Various types of interventions are currently evaluated; however, their appraisal is limited by the lack of reported data concerning the treatment dose and duration. Figure 4 demonstrates the number of trials by the median of planned inclusions per trial for the ten most frequently encountered therapies. Although remdesivir is tested in only 7 trials, these studies have the highest median number of planned inclusions per trial (453, IQR 397-650). On the other hand, cell therapies are assessed by the highest number of trials (n=25), but with a disproportionately low median number of planned inclusions (30, IQR 20-60). Figure 5 shows the total number of planned inclusions and the number of clinical trials for the ten most frequently studied treatments, with hydroxychloroquine being the treatment associated with the highest total number (10,146 planned iclusions).

**Figure 4.**
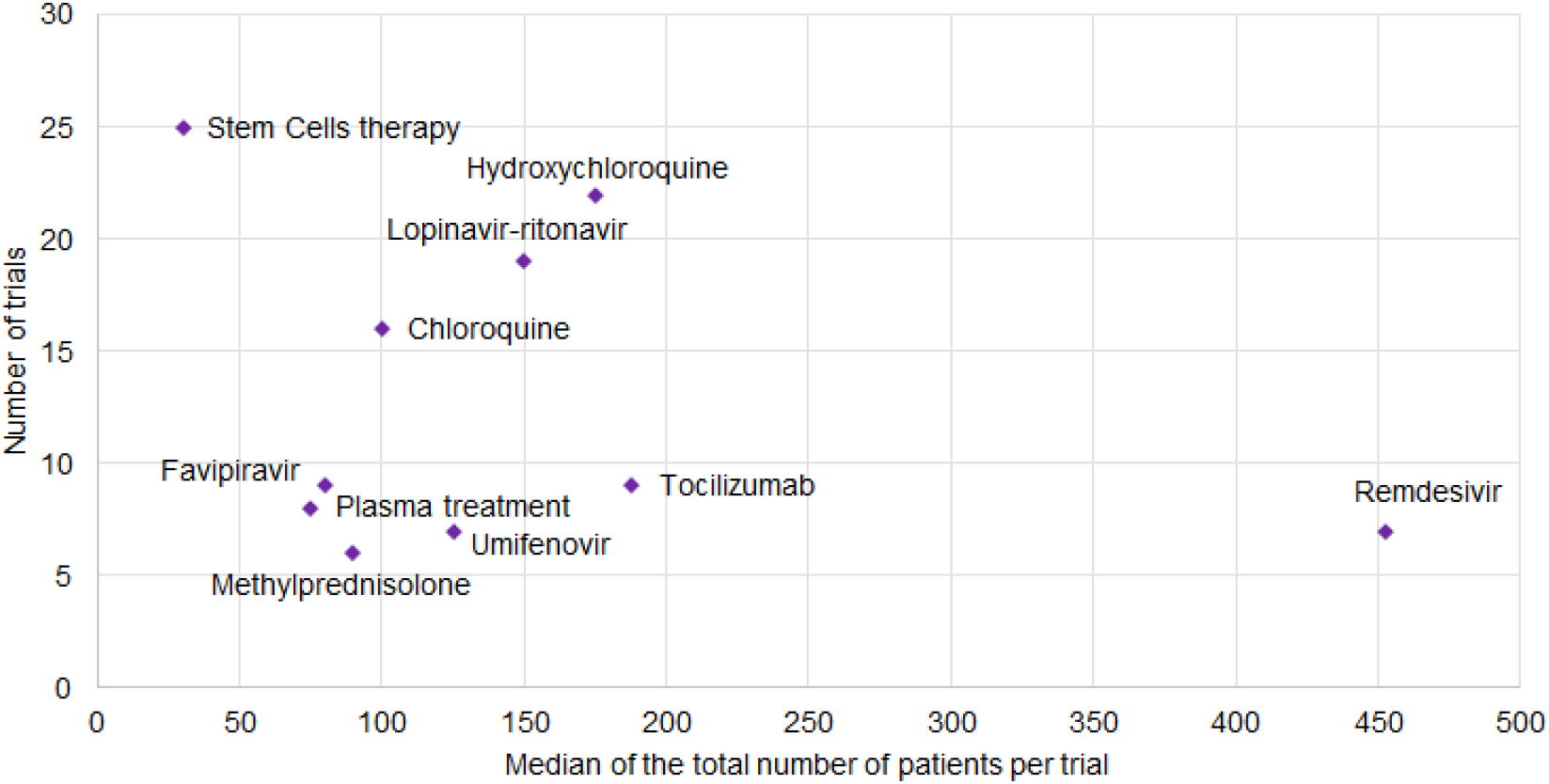
Number of trials reported by the median of the total number of planned inclusions per trial for the most common treatments.

**Figure 5.**
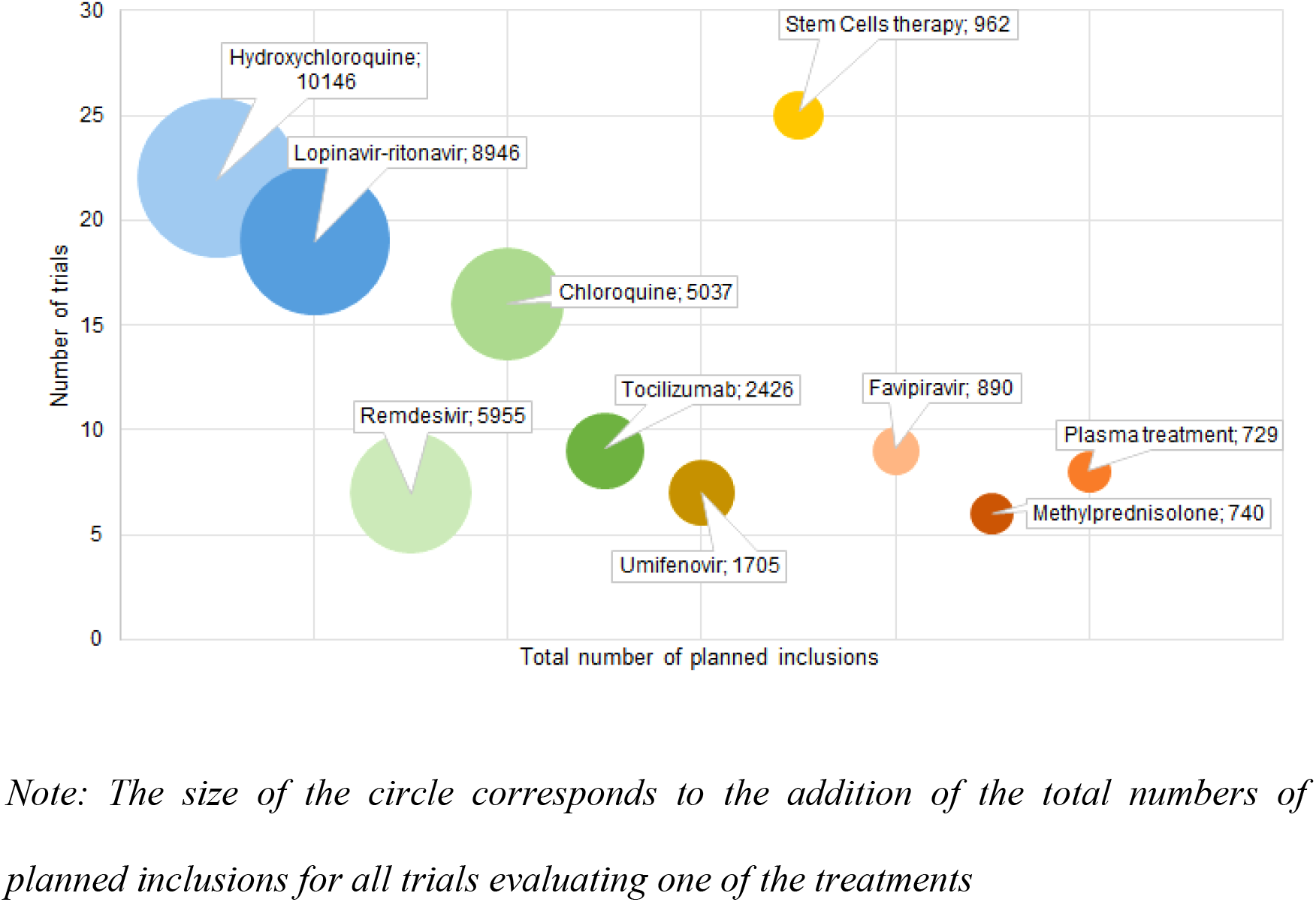
Number of trials per total number of planned inclusions for the ten most frequently assessed treatments.

#### Primary endpoints and outcomes

A clinical primary outcome was defined in 128 therapeutic trials (65%; Table 3); most of them focused on the symptoms’ evolution such as time to clinical recovery, the proportion of patients with clinical improvement or deterioration, length of hospitalisation or mortality. A number of scores are utilized as primary outcomes such as the ordinal 7-point scale adapted from the WHO master protocol, the lung injury score, the pneumonia severity index or the National Early Warning Score 2 score [85–87]. Most prevention studies disclose a clinical primary outcome (13 studies, 81%), while virological, radiological and biological/immunological primary endpoints were reported in 39 (20%), 19 (10%), and 12 (6%) studies respectively (Table 3).

## DISCUSSION

The SARS-CoV-2 is the third emerging coronavirus of the last 20 years, but the one that has led to an evolving pandemic, a fact that urgently requires the implementation of novel therapeutics. However, repurposing of existing drugs is a viable and less time-consuming alternative; licensed broad-spectrum antiviral agents with a well-documented safety profile are currently tested against SARS-CoV-2. Alternatively, screening of chemical compounds’ libraries may be useful in the discovery of an efficacious treatment against COVID-19 disease [8]. In this review, we have summarised and methodologically appraised the ongoing therapeutic and preventive trials for COVID-19.

Therapeutic strategies should follow the two-phased immune response to COVID-19. Initially, treatment should aim at strengthening the host's immune response against the virus, and inhibiting the viral replication [88]. Early initiation of antiviral therapy has been proved beneficial for the prognosis of patients[89]. As advanced age correlates with higher viral loads, older patients could strongly benefit from the prompt initiation of antivirals[90]. Moreover, high inflammatory cytokine levels have been correlated with disease severity and the extentt of lung damage[91]. Thus, in the later phase of the infection, patients could be availed by anti-inflammatory therapeutic approaches such as the Janus kinase inhibitors, blood purification or tocilizumab [92,93].

The Chinese National Health Commission recommends antiviral therapy with the protease inhibitors lopinavir/ritonavir. Other recommended antiviral agents include interferon alpha and chloroquine phosphate [93]. Advantages of chloroquine include its long clinical history, known side effects and low cost, which render it a good candidate for global use[33]. The broad-spectrum antiviral agent remdesivir has successfully been used in individual COVID-19 cases by the Washington Department of Health[94]. Effectiveness of both chloroquine and remdesivir against COVID-19 remains to be seen in ongoing trials.

Prevention is a key player in combating pandemics. The mRNA 1273 vaccine, is currently in phase I, but is not expected to be commercially available this year[95]. The prophylactic potential of hydroxychloroquine/chloroquine is also actively being examined among healthcare professionals[96]. Moreover, prompt testing and isolation are of utmost importance for disease prevention. At the moment, real-time quantitative polymerase chain reaction is the reference standard in diagnostics [97]. The FDA has recently announced the authorisation of rapid molecular tests that are capable of delivering results within minutes, thus facilitating early treatment initiation(when viral loads are highest) and timely isolation[90,98,99].

Based on the retrieved data, cell therapies and hydroxychloroquine were the most frequently evaluated candidate therapies (25 and 22 trials respectively), while remdesivir was associated with the highest median number of planned inclusions per trial (453, IQR 397-650). Although TCMs and homeopathy represent a large proportion of the identified interventional studies, we excluded them from our methodological analysis as we do not have the expertise to analyse clinical trials testing these agents.

This review shows the considerable amount of clinical trials that are currently registered. Although the number of identified trials was high, there were several methodological caveats. Firstly, study design data and details on the interventions being assessed were often lacking. This hampers the available information to researchers and relevant stakeholders, and potentially influences the discovery of successful treatments.

Secondly, most trials, and especially those registered at the beginning of the pandemic, disclosed low participant numbers, which may impact the robustness of their future results. However, these numbers should be cautiously interpreted, as they represent the anticipated, and not the actual number of inclusions. Thirdly, reported primary endpoints were highly heterogeneous among studies. In our opinion, the applicationof clinical primary endpoints should be encouraged in an infection where the association between the clinical status and viral clearance, radiological or immunological evolution is obscure.

The identification of the best therapeutic approach is challenging, especially in the context of a pandemic with thousands of casualties. Our study underlines the need to meticulously register as many study details as possible on international registries during outbreaks, in order to guide the development and enhance the consistency of future trials. The development of clinical trials during an outbreak is an adaptive process and new evidence emerges at an impressive rate. A review of the early registered clinical trials is an important asset for researchers and methodologists. These results might encourage transparency when developing and registering future clinical trials and help improve their methodology, hence the robustness of their results.

In the absence of reliable vaccines and therapies, disease containment plays a pivotal role, as has been recently observed in China. Undertaking actions such as nationwide temperature measurements, health recommendations, detection and strict isolation proved to slow down the transmission rate [100]. The time thereby won is precious for the development of therapies and vaccines. This approach is especially useful for countries with weaker health systems that lack sufficient medical supplies and infrastructure, and often depend on the support of the international community.

## Data Availability

The authors confirm that the data supporting the findings of this study are available within the article [and/or] its supplementary materials.

## Transparency declaration

### Author contributions

Conception and design: PCF, ST, DB, NPS, FM, CL. Acquisition of data: PCF, EK, CDM, HJ, CS, DB, NPS, YY. Interpretation of data: all authors. Drafting the article: PCF, EK, CDM, HJ, CS, DB, NPS, FM, CL, YY. Revising critically the manuscript for important intellectual content: PCF, CS, ST. PCF and DB contributed equally as first authors, CDM and NPS contributed equally as second authors, CL and ST contributed equally as senior authors. All authors approved the final version of the manuscript submitted.

